# Effectiveness of a single dose of oral cholera vaccine: findings from epidemiological and genomic surveillance of *Vibrio Cholerae* in the Democratic Republic of the Congo (PICHA7 Program)

**DOI:** 10.1101/2024.12.16.24318874

**Authors:** Christine Marie George, Alves Namunesha, Kelly Endres, Willy Felicien, Presence Sanvura, Jean-Claude Bisimwa, Jamie Perin, Justin Bengehya, Ghislain Maheshe, Cirhuza Cikomola, Lucien Bisimwa, Alain Mwishingo, David A. Sack, Daryl Domman

## Abstract

This study investigated whole-cell oral cholera vaccine (kOCV) single-dose effectiveness and transmission dynamics of *Vibrio cholerae* through 4 years of epidemiological and genomic surveillance in Democratic Republic of the Congo (DRC). Whole genome sequencing was performed on clinical and water *V. cholerae* strains from 200 patient households and found annual bimodal peaks of *V. cholerae* clade AFR10e. 1154 diarrhea patients were enrolled with 342 culture confirmed cholera patients. A large clonal cholera outbreak occurred 18 months after a kOCV campaign of >1 million doses of Euvichol-Plus, likely because of low vaccine coverage in informal settlements (9%). Clinical and water *V. cholerae* strains in the same household were more closely related than different households suggesting both person-to-person and water-to-person transmission. Single-dose kOCV vaccine effectiveness in the first 24 month after vaccination was 56.9% (95% CI: 18.6%-77.2%), suggesting a single-dose provided modest protection against medically attended cholera during the 24 months post-vaccination.

## INTRODUCTION

An estimated 2.9 million cholera cases and 95,000 deaths occur annually in cholera-endemic countries.^1^ The Democratic Republic of the Congo (DRC) has one of the highest rates of cholera in Africa.^2^ In 2017, the Global Task Force on Cholera Control (GTFCC) released the “*Ending Cholera-A Global Roadmap to 2030”* to reduce cholera deaths by 90% and cholera elimination in 20 additional countries by 2030.^3^ To address cholera in transmission hotspots, the GTFCC recommends oral cholera vaccine (OCV) campaigns and case-area-targeted-interventions (CATIs) of water, sanitation, and hygiene (WASH) around cholera cases. However, there is currently an OCV stockpile shortage globally resulting in the need for evidence on the duration of protection conferred by OCV to determine the optimal frequency to repeat OCV campaigns.^4^ Most studies on OCV effectiveness are from India and Bangladesh, and evaluate the effectiveness of two-dose OCV protection.^5^ Only four studies have evaluated single-dose OCV vaccine effectiveness (with only two for a duration of 24 months or longer), and all except one used the no longer produced Shanchol vaccine.^5^

Few studies combine evaluations of OCV effectiveness with genomic surveillance of clinical and environmental *Vibrio cholerae* strains using whole genome sequencing (WGS) which can provide valuable information on how OCV campaigns impact *V. cholerae* transmission dynamics and the genetic characteristics of *V. cholerae* strains circulating. WGS of *V. cholerae* strains also presents a valuable tool to link epidemics globally and investigate cholera transmission dynamics through distinguishing strains based on single nucleotide polymorphisms (SNPs). Genomic data from 45 African countries revealed that seventh pandemic El Tor (7PET) *V. cholerae* strain was introduced into Africa at least 15 times since 1970.^8^ Previous genomic studies (2018-2024) in DRC found *V. cholerae* 7PET strains to be in the AFR10d, AFR10e clade, and ARFR10w clade sublineages.^9,10^

Only one study has performed WGS on water source and clinical *V. cholerae* strains from patient households to investigate cholera transmission dynamics.^11^ This study conducted by our research group in Bangladesh found that 80% of cholera patient households had source water strains closely related to clinical strains, and 20% of households had clinical strains from infected individuals that were more closely related to clinical strains from another household than to source water strains from their own household. These results were consistent with person-to-person and water-to-person *V. cholerae* transmission. There have been no genomic studies to elucidate transmission dynamics of *V. cholerae* infection for cholera patient households in sub-Saharan Africa, with previous studies exclusively in South Asia.

In this study we conducted 4 years of epidemiological and genomic surveillance of *V. cholerae* in eastern DRC to achieve the following study objectives: (1) to evaluate the effectiveness of a single-dose of killed whole-cell oral (kOCV) Euvichol-Plus during the 24 month period after a preventative kOCV campaign; (2) to investigate *V. cholerae* transmission dynamics among cholera patients, household members, and water sources using WGS; and (3) to investigate the spatiotemporal spread of *V. cholerae* in this hyper-endemic region.

## METHODS

### Ethical approval

This study was conducted in urban Bukavu in South Kivu province, DRC. We received ethical approval for this study from the Johns Hopkins Bloomberg School of Public Health and Catholic University of Bukavu. All participants or their guardians gave written informed consent.

### Study Design and Protocol

For cholera surveillance activities (passive surveillance), diarrhea patients admitted to 115 healthcare facilities in Bukavu from March 2020-March 2024 had whole stool tested for *V. cholerae* by bacterial culture. Cholera patients were defined as diarrhea patients with a stool bacterial culture result positive for *V. cholerae*. A prospective cohort study of household contacts of cholera patients was conducted from December 2021-December 2023 to investigate cholera transmission dynamics in cholera patient households. Household contacts of cholera patients were defined as those sharing a cooking pot and residing in the same home with the cholera patient for the past three days. Household contacts were enrolled within 24 hours of cholera patients. The sample size for the prospective cohort study was determined by the number of cholera patients that could be screened and were willing to participate in the cohort study from December 2021-December 2023. Cholera patient households in the prospective cohort study were visited on Days 1, 3, 5, 7 and 9 (Visits 1-5) after the household’s index cholera patient was admitted at a health facility to conduct clinical surveillance. For clinical surveillance, a stool sample was collected from household contacts at each household visit to test for *V*. *cholerae* by bacterial culture. An unannounced spot check was conducted at each timepoint to collect a sample of the household’s source and stored drinking water to test for *V*. *cholerae* by bacterial culture.

During December 28^th^, 2021-January 2^nd^, 2022 and March 31^st^-April 4^th^, 2022, a preventative OCV campaign delivered 1.04 million doses of Euvichol-Plus kOCV in Bukavu. This campaign was delivered through a combination of door-to-door visits and designated healthcare facilities. The kOCV vaccination status for diarrhea patients and their household members was assessed through self-report or caregiver-report during the time of patient treatment in the healthcare facility or during a home visit conducted the same or the following day. A structured questionnaire was administered obtaining information on kOCV administration and the date and number of doses. A kOCV vaccine card was shown along with a photo of a person consuming kOCV Euvichol-Plus. Informal settlements were defined as areas where there were no household connections to piped water.

### Laboratory Analyses

All whole stool samples were brought to the PICHA7 Enteric Disease Microbiology Laboratory within three hours of the sample being produced, and water samples were brought to the laboratory within 3 hours of collection for *V*. *cholerae* analysis by bacterial culture according to our previous methods.^12^ Strains were then preserved as stabs in nutrient agar or on Whatman filter paper to preserve their DNA.

### Whole genome sequencing

Genomic DNA for strains on filter paper was extracted using published Chelex methods and the ZymoBIOMICS DNA Miniprep Kit was used for strains preserved as stabs.^13,14^ WGS was performed at the SeqCenter in Pittsburgh, Pennsylvania and the University of New Mexico Health Sciences Center. Short reads were processed using the Bactopia pipeline^15^ and SPAdes v.3.10.0^16,17^ and annotated using Prokka v.1.520.^18^ Genome completeness estimates and checks for contamination were performed using CheckM v.1.0.722 and Kraken v.0.10.6.^19,20^

### Genomic and phylogenetic analyses

Paired-end reads for 255 7PET strains were mapped and variants called against the *V. cholerae* O1 El Tor reference N16961 (NCBI accession numbers LT907989 and LT907990) using snippy v.4.6.0 (https://github.com/tseemann/snippy) via the Bactopia pipeline^15^ to generate a reference-based alignment containing 329 variable SNP sites. All 255 strains mapped over 90% of the reference genome and were considered for further analysis. A pairwise SNP matrix was generated for the 255 7PET strains in this study using ‘pairsnp’ (https://github.com/gtonkinhill/pairsnp). We used ARIBA v.2.14.7 to determine mutations in *wbeT* using a reference wild type Ogawa *wbeT* gene (ENA accession AEN80191.1).

All 7PET strains from this study were combined with 1,418 globally representative 7PET strains to assess phylogeographic relatedness.^21^ The 12,561 variable site (SNP) alignment was used to build a maximum likelihood phylogeny using IQ-Tree v1.6.12 under the GTR model with the gamma distribution to model site heterogeneity (GTR+GAMMA) with 10,000 ultrafast bootstraps and 10,000 bootstraps for the SH-like approximate likelihood ratio tests (SH-aLRT).^22^ The phylogeny was visualized with ggtree v.1.6.11^23^ and rooted using the pre-seventh pandemic strain M66.

### Local transmission analyses

Insights on local transmission dynamics were gleaned from a 329 SNP reference-based alignment of the 255 7PET strains. A maximum-likelihood tree was built using IQ-Tree under the GTR+GAMMA model with 10,000 ultrafast bootstraps and 10,000 SH-aLRT. Phylogenies were visualized in Microreact.^24^ Minimum spanning trees were created using *GrapeTree* v.1.5.0.^25^ Geospatial data for cases was visualized using python with the *geopandas*^26^ and *contextily*^27^ packages.

### Data availability

All next-generation sequencing data generated in this study have been deposited into the NCBI Sequence Read Archive (SRA; https://www.ncbi.nlm.nih.gov/sra).

### Statistical Analysis

To assess single dose Euvichol-Plus kOCV vaccine effectiveness during the 24-month period after vaccination using a test-negative design among diarrhea patients, logistic regression models were performed with cholera infection as the outcome (defined as a positive bacterial culture result for *V. cholerae*) and the predictor was kOCV vaccination status (whether diarrhea patients reported receiving one dose of kOCV during the dates of the kOCV campaign. In this model the odds ratio (OR) was the odds of single-dose kOCV vaccination in the cases compared to controls. Vaccine effectiveness was calculated as (1-OR x 100%), and kOCV vaccinated individuals reporting receiving two doses of kOCV were excluded from the analysis. This analysis was performed in SAS (version 9.4). Permutation tests using R and python were performed to analyze pairwise comparisons of genomic data. For pairwise comparisons, SNP counts for each strain were compared to the reference strain reference N16961.

## RESULTS

### Epidemiology

During the 4 years of epidemiological and genomic surveillance from March 2020 to March 2024, 342 out of 1154 diarrhea patients (30%) were identified to be *V. cholerae* by bacterial culture (Figure 1). Figure 2 shows a map of the 115 surveillance healthcare facilities and diarrhea patient households. An annual bimodal peak of cholera was observed corresponding with the dry season (June to August) and the rainy season (September to January) in 2020 and 2023. In December 2021, an kOCV campaign distributed 1.04 million doses of Euvichol-Plus within Bukavu (study surveillance site). After this kOCV campaign there were sporadic cholera patients through January 2023. Then, 18 months after the kOCV campaign from June to November of 2023, there was a large cholera outbreak in this same area. Nine percent (309/3395) of individuals in the informal settlements in our study area reported receiving at least one kOCV dose during our vaccine coverage surveys between December 2021 and April 2022. During the four-year surveillance period, stored and source water samples were collected from 177 cholera patient households with 9% (16/172) of households with stored water samples positive by bacterial culture for *V. cholerae* and 5% (8/151) of households with positive source water samples. In total 29 water samples were *V. cholerae* positive by bacterial culture over the study period (Supplemental Figure 1). From October 2021 to November 2022 (including the 12-month period after the kOCV campaign) there was no *V.cholerae* detected in drinking water samples despite there being culture confirmed cholera patients.

**Figure 1.**
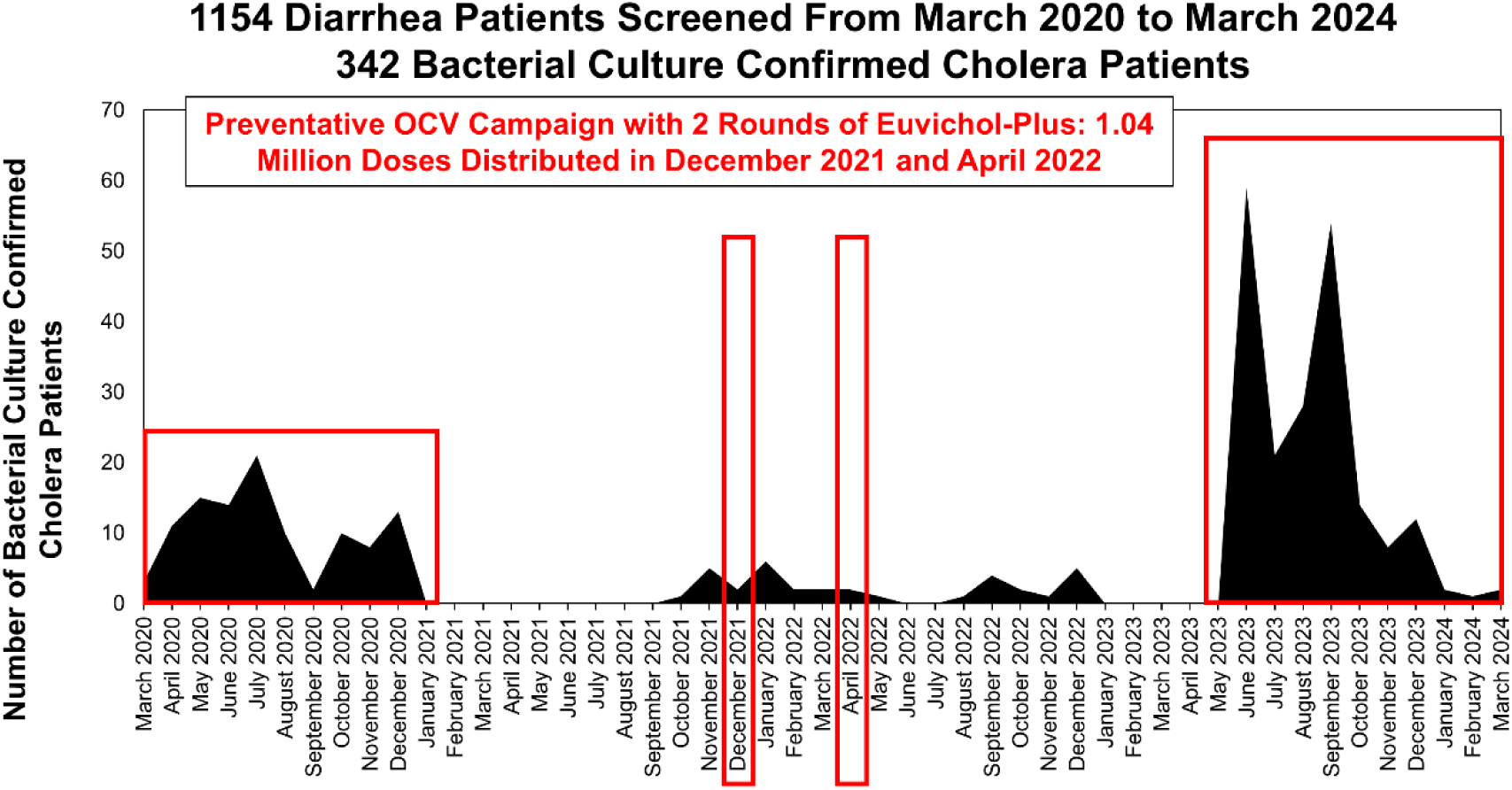
Four-Year Epidemological Surveillance of Bacterial Culture Confirmed Cholera Patients from 115 Health Facilities in Bukavu City of South Kivu Province of the Democratic Republic of the Congo from March 2020 to March 2024. A total of 1154 diarrhea patients were screened at 115 health facilities and 342 cholera patients were confirmed by bacterial culture. A preventative oral killed whole-cell cholera vaccine (kOCV) campaign was conducted in December 2021 and April 2022.

**Figure 2.**
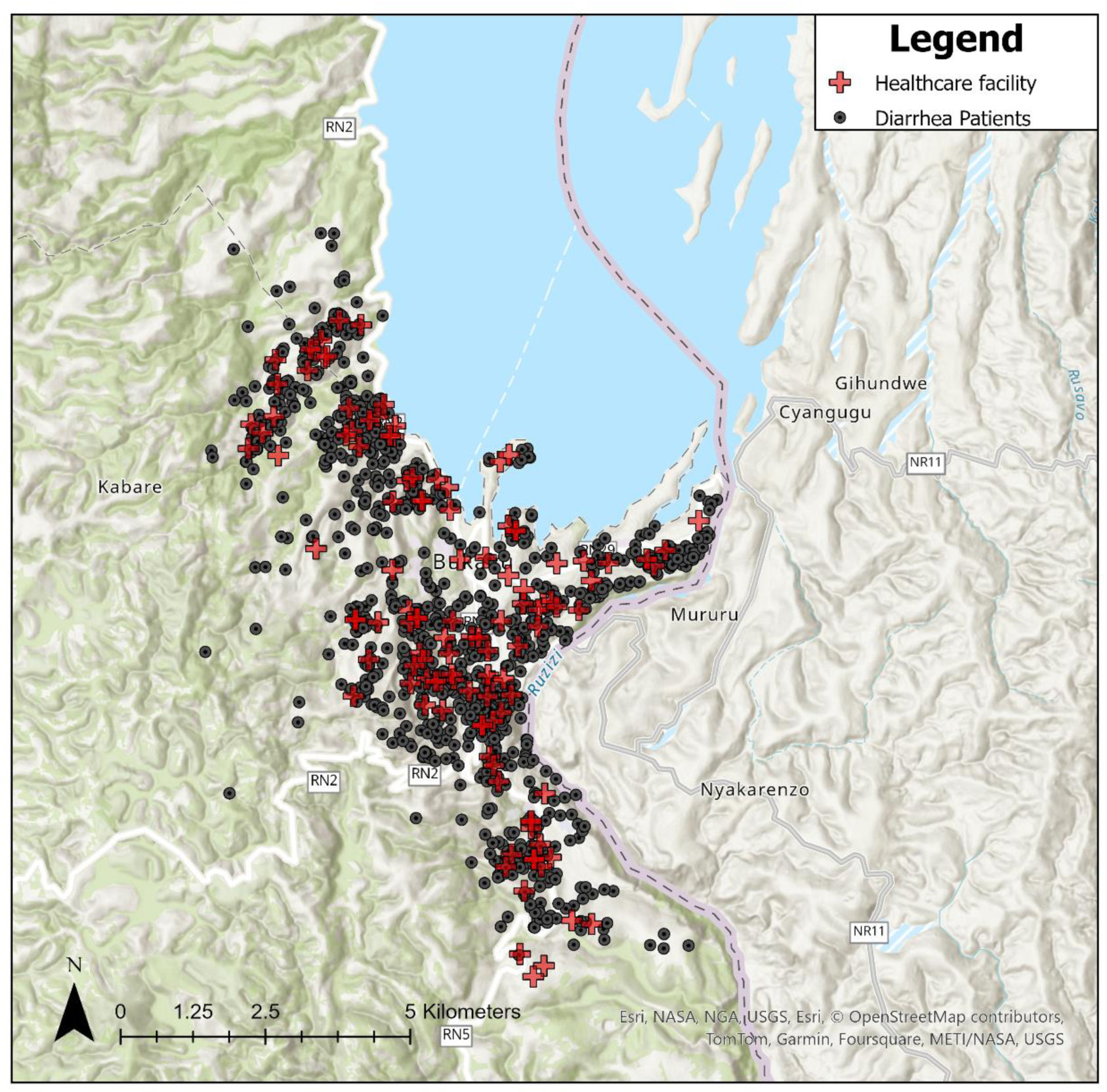
Map of diarrhea patients positive for V. cholerae by bacterial culture and health facilities where cholera patient surveillance was conducted between 2020-2024 in Bukavu, Democratic Republic of the Congo. Red cross indicates healthcare facilities (N=115) and black dots indicate diarrhea patient households (N=1098) (56 patient households were missing GPS coordinates).

### OCV Vaccine Effectiveness

Seven-hundred-and-fifty diarrhea patients were admitted to surveillance healthcare facilities during the 24-month period after the kOCV campaign (December 2021 to December 2023). Demographic characteristics for kOCV vs. non-kOCV vaccinated individuals are included in Table 1. Ninety-four percent (708/750) of diarrhea patients reported residing in their current residence for the three nights before hospitalization, and 12% had running water inside the home (93/748). Twelve percent (93/750) (15 cholera patients and 78 non-cholera diarrhea patients) of diarrhea patients reported receiving at least one dose of kOCV from December 2021 to April 2022, with only 2% (14/750) of patients reporting receiving two kOCV doses. Thirteen percent of kOCV vaccinated patients (12/93) showed a vaccination card. During this period there were 208 cholera patients (193 non-kOCV vaccinated and 15 kOCV vaccinated). Five-hundred-and-thirty-one diarrhea patients during this 24-month period were >1 years of age. The unadjusted single-dose kOCV vaccine effectiveness in the first 24 month after vaccination was 59.8% (95% CI: 19.7% to 79.9%) for individuals >1 year and after adjustment for age (continuous) the vaccine effectiveness was 58.7% (95% CI: 17.3% to 79.4%). Ten diarrhea patients were excluded from this analysis because they received 2 doses of kOCV. Supplementary Table 1 includes the single-dose kOCV vaccine effectiveness by time interval for individuals >1 year.

**Table 1.** Diarrhea patient demographic characteristics by OCV status and cholera status.

| All diarrhea patients |  |  |  | OCV status |  |  |  |  |  | P-value |
| --- | --- | --- | --- | --- | --- | --- | --- | --- | --- | --- |
|  |  |  |  | Received OCV |  |  | No OCV |  |  |  |
|  | % | n | N | % | n | N | % | n | N |  |
| Diarrhea Patients |  |  | 750 |  | 93 | 750 |  | 657 | 750 |  |
| Age |  |  |  |  |  |  |  |  |  |  |
| Median ± SD (Min - Max) (Years) |  | 5 ± 19 (0-82) |  | 2 ± 15 (0-63) |  |  | 7 ± 19 (0-82) |  |  |  |
| 0-1 Years | 29% | 217 | 750 | 32% | 30 | 93 | 28% | 187 | 657 | 0.527 |
| 1-4 Years | 20% | 148 | 750 | 30% | 28 | 93 | 18% | 120 | 657 | 0.011 |
| 5-14 Years | 13% | 96 | 750 | 12% | 11 | 93 | 13% | 85 | 657 | 0.893 |
| 14 Years or Greater | 39% | 289 | 750 | 26% | 24 | 93 | 40% | 265 | 657 | 0.009 |
| Gender |  |  |  |  |  |  |  |  |  |  |
| Female | 49% | 368 | 750 | 48% | 45 | 93 | 49% | 323 | 657 | 0.977 |
| Running water inside home | 12% | 93 | 748 | 13% | 12 | 93 | 12% | 81 | 655 | 1.000 |
| Working mobile phone in home | 85% | 565 | 663 | 92% | 78 | 85 | 83% | 487 | 587 | 0.097 |
| Individuals residing in home |  |  |  |  |  |  |  |  |  |  |
| Median ± SD (Min - Max) (Years) |  | 7 ± 3 (1-23) |  | 7 ± 2 (3-16) |  |  | 7 ± 3 (1-23) |  |  | 0.098 |
| Years of formal education* |  |  |  |  |  |  |  |  |  |  |
| Median ± SD (Min - Max) (Years) |  | 9 ± 5 (0-23) |  | 7 ± 5 (0-15) |  |  | 9 ± 5 (0-23) |  |  | 0.028 |

SD=Standard Deviation
\* for those age 18 and over

### Genomic analyses

We sequenced 255 *V. cholerae* 7PET genomes collected between December 2020 to December 2023 from the study area described above. Of these strains, 247 were clinical (158 Ogawa and 89 Inaba) strains representing 243 individuals from 200 households (198 patients and 45 household contacts), and 8 were water samples. The WGS analysis included 75% of cholera patient households (255/342) during the surveillance period, all strains with isolates available were sequenced. To investigate the genetic relatedness of the *V. cholerae* strains, we analyzed the pairwise SNP differences across the 255 strains. Of the 200 households in the study, 31 households are represented by more than one sample, 26 households had more than one participant with samples, and five households had both a clinical and water sample. Figures 3A and 3B show the maximum-likelihood phylogeny of the 255 7PET *V. cholerae* genomes by year, sample type, and serotype. Our phylogenic analyses indicate *V. cholerae* strains isolated from both clinical and water samples were closely related, indicating limited genetic divergence over the study period. Figure 4A shows a violin-plot of pairwise differences between all *V. cholerae* strains, those from the same households, and those from pre- and post-vaccine campaign.

**Figure 3.**
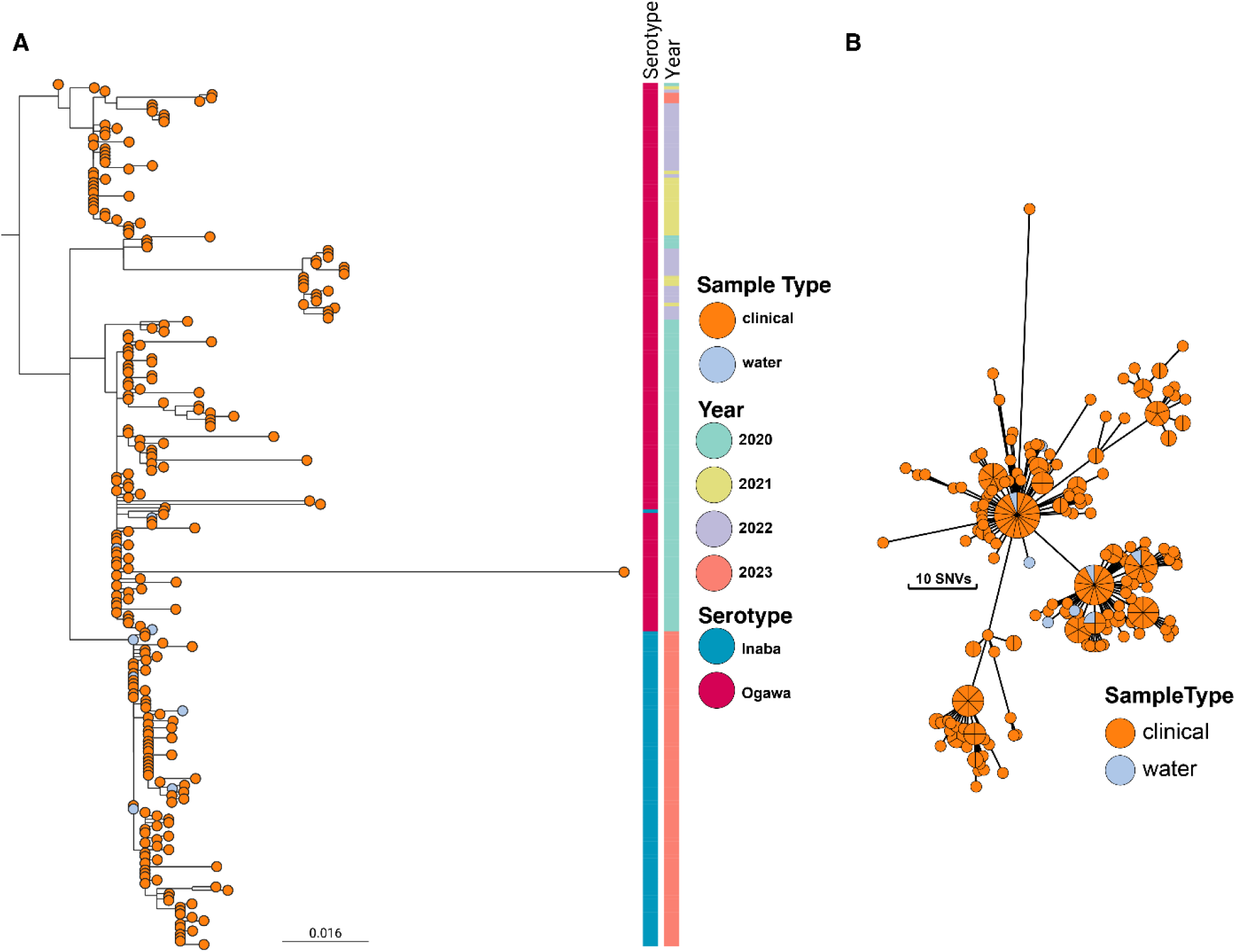
Phylogenetic analyses describing the relationships of the 255 7PET strains. **A.** Maximum likelihood phylogeny of the 255 7PET *V. cholerae* genomes sampled in Bukavu from 2020 to 2023. The nodes are colored according to sample type. The associated colored metadata corresponds to year of sampling and inferred serotyping results based on genome analysis. Not all strains have serotype data as indicated. **B.** Minimum spanning tree of 255 genomes. Strains that have zero SNP differences between them are collapsed into a single node. Node sizes scale with number of samples. The nodes are colored according to sample type.

**Figure 4.**
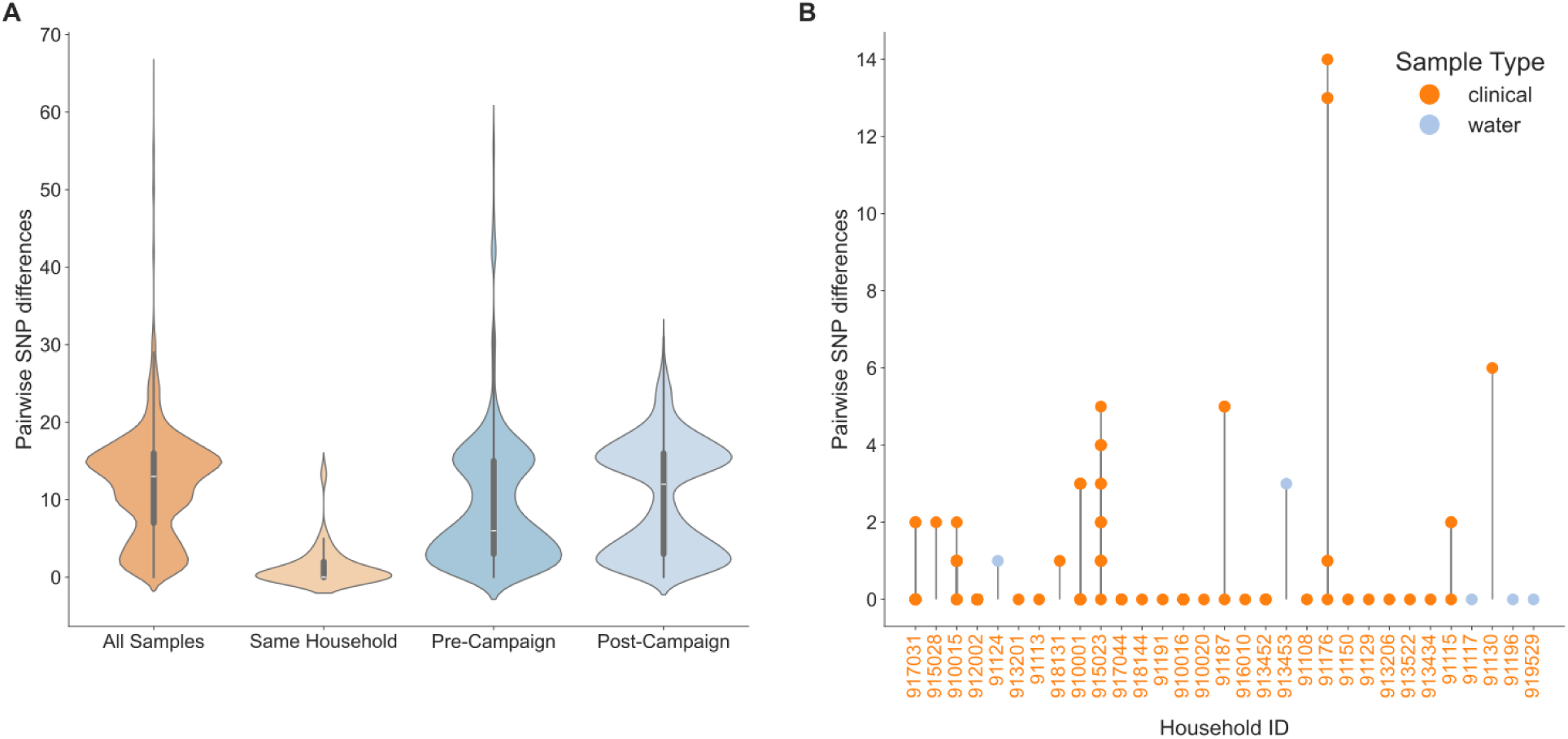
Pairwise comparisons of SNPs. (A) Boxplots of pairwise SNP differences. Boxplots depict the distribution of pairwise SNP differences for all strains, those from the same household, and those from the same individual. Each plot shows the median, and interquartile range. The number of pairwise comparisons for each category is as follows: All samples (n=32,385), same household (n=99), Pre kOCV Campaign (n=6,205), and Post-kOCV Campaign (n=10,296). **(B) Stem plots of the SNP differences per household**. Pairwise SNP differences are relative to the first sample in the household. Nodes are colored by sample type and stems connect all samples to their given household.

Between all strains, the minimum number of SNP differences was 0 and the maximum number was 65, with a median of 13. Of strains from the same household, the range was from 0-14 SNPs, with a median of 0 SNPs (Figure 4B). We found a significant difference in the median number of SNPs between strains from different households and those from the same household (p=<0.0001; Mann-Whitney U test). For strains from the same individual, the range was 0-1 SNP, with a median of 0 SNPs. Five households had both a water sample and a clinical sample, where in three households these strains had zero SNP differences between them. The other two households had 3 and 1 SNP differences between the clinical and water samples. This provides strong evidence that the same strain causing infections in the household is also detected in the household water. However, we cannot determine whether the water was the source of infection, or the water was contaminated after an infection was present in the household. The median difference in the number of SNPs was significantly lower for strains from cholera patients pre-kOCV vaccine campaign (median: 6 range: 0-58) compared to post-kOCV vaccine campaign (median: 12 range: 0-31) (Mann-Whitney U test; P-value: p<0.0001)).

To understand how the 255 *V. cholerae* strains from this study fit within the larger diversity of 7PET strains, we contextualized these strains within a collection of 1,422 additional 7PET genomes (Figure 5).^21^ Our phylogenetic analysis placed these strains within the T10 lineage within the AFR10e clade. Our strains, sampled from 2020-2023, were similar to other recent samples (2015-2020) from the DRC in the same region of South Kivu.^10^ Our genomic analysis of the *wbeT* gene indicated that samples from 2020-2022 have a wildtype, intact gene which would indicate an Ogawa serotype. There is a single exception for a strain sampled in 2020 that had a frameshift mutation (N165fs), which would likely result in an Inaba serotype. All samples in 2023 after the OCV campaign, however, appear to have a fragmented *wbeT* gene marked by insertion of mobile elements into this gene – likely conferring an Inaba serotype. In line with other AFR10e strains, our strains harbor mutations in *gyrA* (S83I) and *parC* (S85L) which have been previously described as resulting in reduced susceptibility to fluoroquinolones.^10^

**Figure 5.**
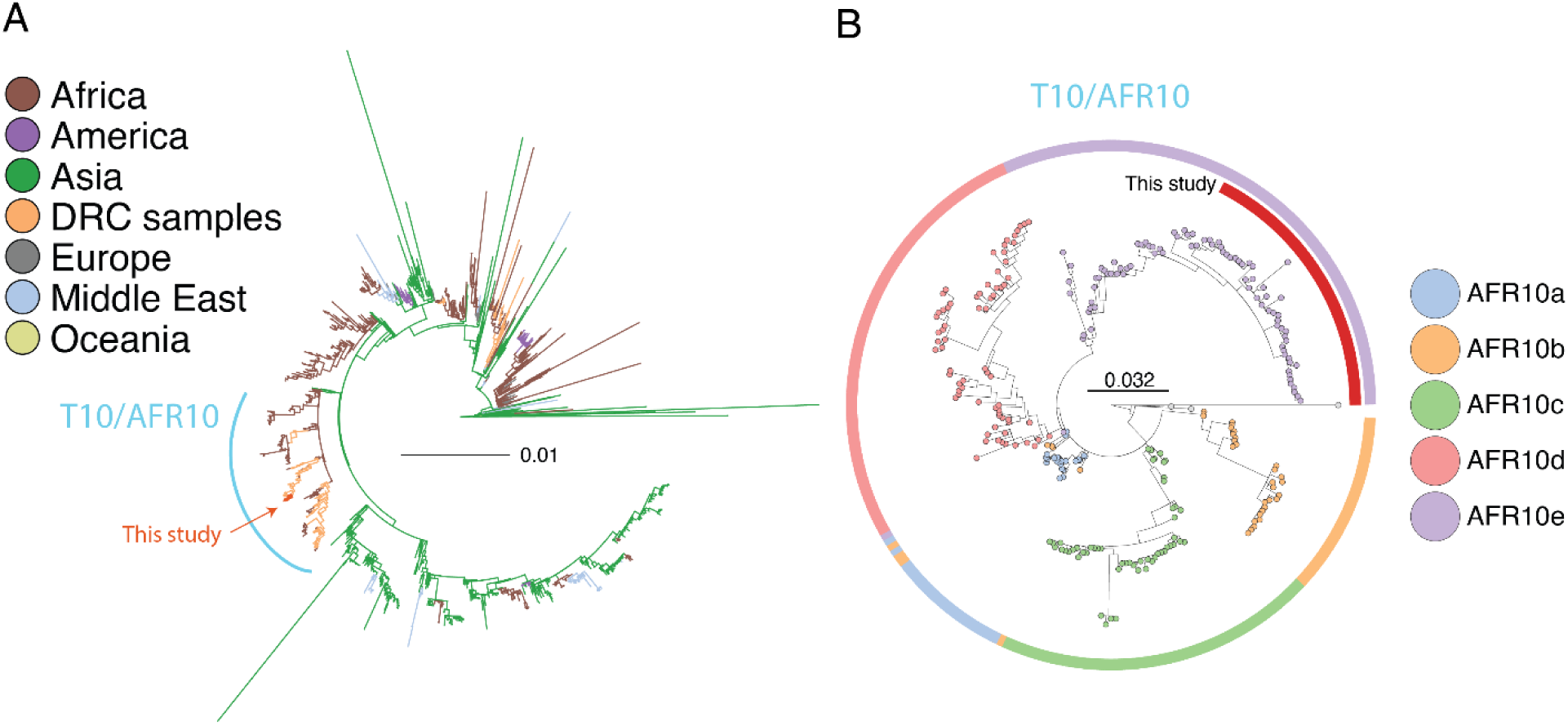
Maximum likelihood phylogenetic trees of seventh pandemic strains. (**A) Globally representative phylogeny of 1,428 7PET strains.** Five representative strains from this DRC study were placed within the larger context of 1,423 7PET strains. The tree was rooted on A6 strain. The branches are colored according to geographic origin of the strain. **(B) Phylogeny of the T10/AFR10 lineage.** Representative strains from this DRC study (n=46) were placed within the context of 221 T10/AFR10 lineage strains. The tree was rooted on the reference strain N16961.

## DISCUSSION

In our urban cholera endemic setting there were annual bimodal peaks of the *V. cholerae* clade AFR10e corresponding with the dry and rainy seasons. A third of diarrhea patients presenting at our 115 surveillance healthcare facilities were confirmed to have cholera by bacterial culture. Nine percent of stored water and five percent of source water samples from cholera patient households had *V. cholerae* present. This finding suggests that both source water and contamination of stored water may be potential transmission routes for *V. cholerae* infections in cholera patient households. A large clonal cholera outbreak was observed 18 months after a single-dose kOCV campaign where >1 million doses of Euvichol-Plus were distributed. This large outbreak occurred in spite of the kOCV vaccine campaign, perhaps because of the low kOCV coverage (9%) in the informal settlements in Bukavu, which are often hotspot areas for cholera because of limited access to improved drinking water sources and sanitation options and poor hygiene conditions.^28^ There were no water samples positive for *V. cholerae* during the 12-month period after the kOCV campaign, despite the presence of cholera patients. Future studies should explore this finding further by investigating the impact of kOCV campaigns on the persistence of *V. cholerae* in the environment.

Using a test-negative design, we found that a single-dose of Euvichol-Plus provided modest protection against medically attended cholera during the 24 months after vaccination, consistent with previous studies.^5^ This test-negative design to evaluate kOCV effectiveness could be incorporated into existing cholera surveillance activities implemented globally and integrated as part kOCV vaccine campaign rollouts. Our study findings suggests that with increased vaccination coverage in informal settlements, single-dose kOCV campaigns present a promising approach to deliver along with water, sanitation, and hygiene programs to reduce cholera in this setting in DRC.

Our genomic findings highlight several important points. Firstly, we did not detect any samples that grouped into clade AFR10d, a finding that seems to further corroborate that clade AFR10e replaced AFR10d around 2018 in this region.^29^ Second, during the subsequent cholera outbreaks after the large kOCV campaign we observed a change in strain serotype to Inaba which was likely related to a fragmented *wbeT* gene marked by insertion of mobile elements. This is consistent with another study that observed a serotype switch following a vaccination campaign.^6^ Additionally, we observed an increase in the number of SNPs between *V. cholerae* strains from pre to post kOCV campaign. However, both our current findings and the one from the previous study are observational and causality cannot be inferred. Third, this study provided evidence that household water contained the same strains that caused cholera infections among household members. This result combined with the finding that clinical strains in the same household were more closely related than strains from different households, even when no *V. cholerae* was found in household drinking water, suggests a combination of person-to-person and water-to-person cholera transmission. This is the first study, to our knowledge, to conduct both clinical and environmental surveillance of *Vibrio cholerae* in cholera patient households in a sub-Saharan Africa setting. These findings suggest that it is critical that WASH programs put emphasis on both water treatment of household water and handwashing with soap to prevent cholera transmission.

This study has several strengths. The first is the 4-year duration of epidemiological and genomic surveillance of cholera patients and their water sources during a time before and after a kOCV campaign. Previous longitudinal studies including both clinical and environmental surveillance have been almost exclusively in South Asia. The second strength is evaluating the effectiveness of a single-dose of Euvichol-Plus over a 24-month period building on previous studies which focus on two-doses of kOCV and evaluated the Shanchol kOCV which is no longer produced. The only study we are aware of that evaluated the single-dose kOCV effectiveness of Euvichol-plus for a period of 24 months or longer was from DRC which found a single-dose kOCV vaccine effectiveness of 54% from 12 to 17 months after vaccination, similar to our study.^30^ Additionally, the test-negative design for kOCV effectiveness builds on observational studies utilizing community control by reducing the likelihood of differences between cases and control in care-seeking behavior which can introduce substantial bias in observational studies of vaccine effectiveness. The final study strength is the inclusion of the genomics data which allowed us to investigate how water and clinical *V. cholerae* strains were evolving over time during the period before and after a kOCV campaign and to investigate *V. cholerae* transmission dynamics in cholera patient households.

This study also has some limitations. First, our surveillance focused only on an urban setting so we cannot generalize our findings to rural settings. Additional evidence is needed on single-dose kOCV vaccine effectiveness in rural settings globally. Second, as with all observational study design there is the possibility of misclassification of the patient’s vaccination status and cholera infection outcome. Also, it should be noted that the vaccine campaign was a preventive campaign in an area identified as a hotspot; however, currently, because of the global kOCV vaccine shortage, all campaigns are reactive and respond to outbreaks. Future studies are needed to evaluate single-dose kOCV effectiveness during reactive kOCV campaigns globally.

In conclusion, there were annual bimodal peaks of the clade AFR10e of *V. cholerae* in our urban cholera endemic setting in eastern DRC. A third of diarrhea patients admitted to surveillance healthcare facilities were confirmed to have cholera by bacterial culture. A large clonal cholera outbreak occurred 18 months after a single dose kOCV campaign, and the severity of the outbreak may have been related to the low vaccine coverage in informal settlements. Genomics findings suggest both person-to-person and water-to-person transmission. A single-dose of Euvichol-Plus provided modest protection against medically attended cholera during the 24 months after vaccination. These results contribute to the evidence base supporting the use of single dose kOCV campaigns in combination with WASH programs to reduce cholera in endemic settings globally.

## Data Availability

All data produced in the present study are available upon reasonable request to the authors.

## Acknowledgements

Many thanks to the study participants for their support in implementing this study, as well as our funder. Thank you also to our research officers who played a crucial role in the success of this work: Raissa Boroto, Freddy Mwambusa, Pacifique Kitumaini, Blessing Muderhwa, Jean Claude Lunyelunye, Feza Rugusha, Gisele Kasanziki, Brigitte Munyerenkana, Jessy Tumsifu, Pascal Kitumaini, Emmanuel Buhendwa and Julienne Rushago.

This work was made possible with funding from Wellcome and UK aid from the Foreign and Commonwealth Development Office grant number 215674Z19Z and 1R01AI148332-01 provided to Christine Marie George at Johns Hopkins School of Public Health. The views expressed do not necessarily reflect FCDO’s official policies or views. Daryl Domman was supported by supported by an award from the National Institutes of Health (NIH) under grant number UL1TR001449 and/or KL2TR001448. David Sack was supported by 2R01AI123422-06A1. The funding agencies had no involvement in study design, data collection, data analysis, and data interpretation.

Dr. George is an infectious disease epidemiologist and environmental engineer. Her research focuses on developing and evaluating community and healthcare facility–based water, sanitation, and hygiene interventions to reduce infections in low- and middle-income countries and low-resource settings globally.

---

**Supplementay Figure 1.**
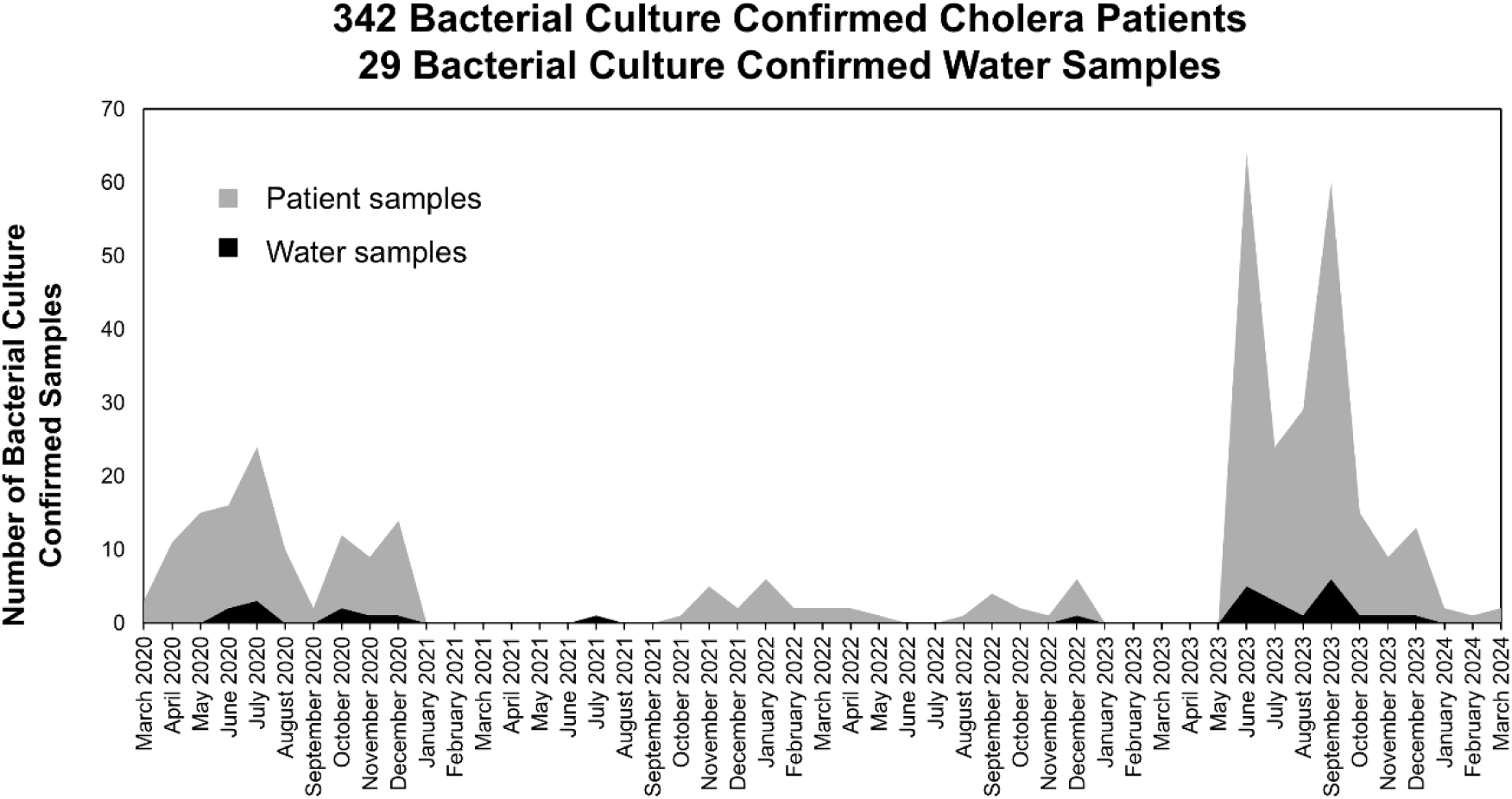
Four-Year Epidemological Surveillance of *Vibrio Cholerae* Bacterial Culture Confirmed Cholera Patients and Water Samples Bukavu City of South Kivu Province of the Democratic Republic of the Congo from March 2020 to March 2024. A total of 1154 diarrhea patients were screened at 115 health facilities and 342 cholera patients were confirmed by bacterial culture. 29 water samples were positive for cholera (9 source and 20 stored).

**Supplementary Figure 2.**
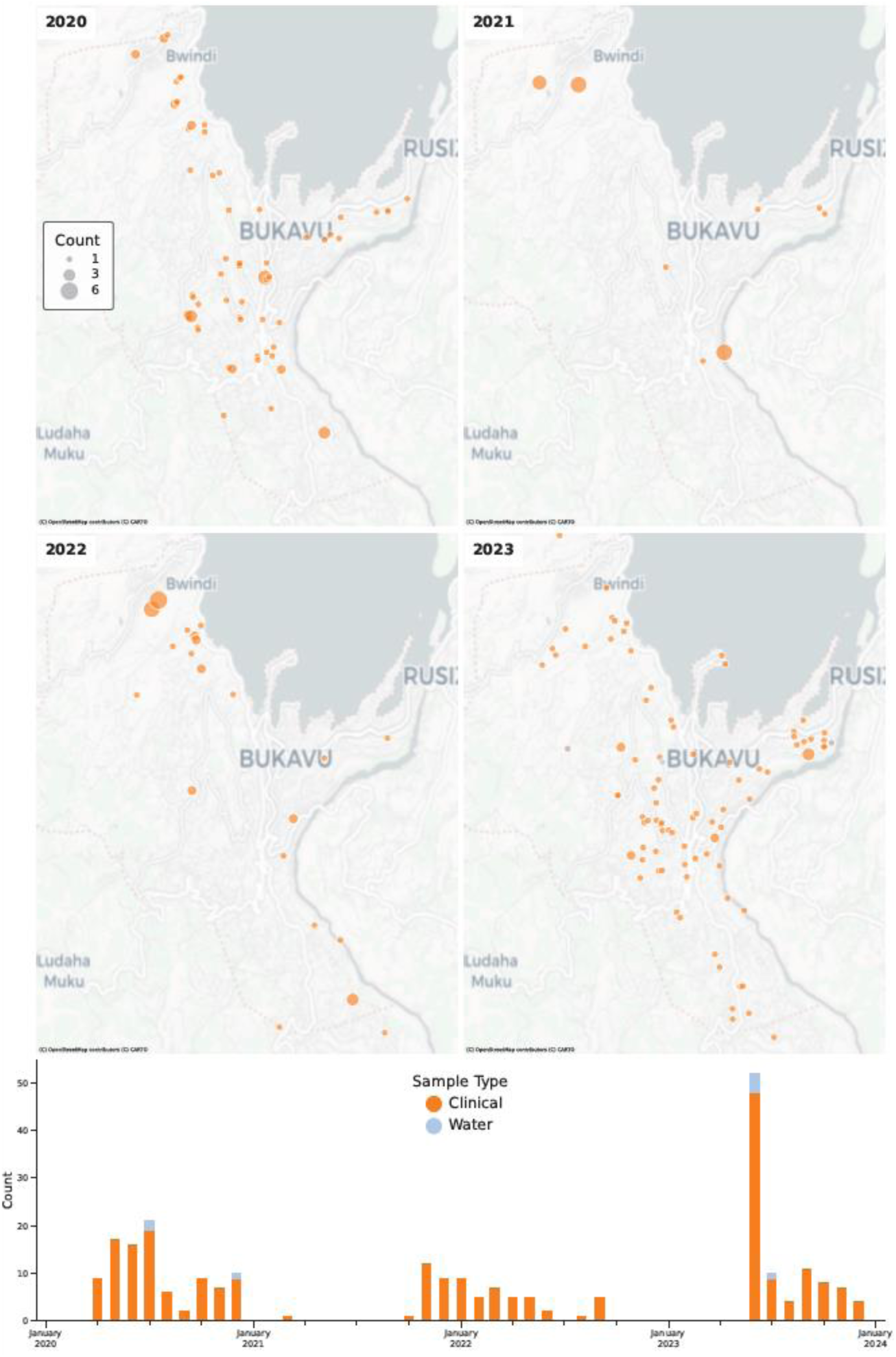
Case counts and locations of sampling for genomic analysis in Bukavu, Democratic Republic of the Congo between 2020 - 2023. Top, map indicates spatial distribution of strains and the sample type per year. Bottom, the number and sample type of strains per month.

